# *PixelPrint^4D^*: A 3D printing method of fabricating patient-specific deformable CT phantoms for respiratory motion applications

**DOI:** 10.1101/2024.08.02.24311385

**Authors:** Jessica Y. Im, Neghemi Micah, Amy E. Perkins, Kai Mei, Michael Geagan, Peter B. Noël

## Abstract

All in-vivo medical imaging is impacted by patient motion, especially respiratory motion, which has a significant influence on clinical workflows in diagnostic imaging and radiation therapy. Many technologies such as motion artifact reduction and tumor tracking algorithms have been developed to compensate for respiratory motion during imaging. To assess these technologies, respiratory motion phantoms (RMPs) are required as preclinical testing environments, for instance, in computed tomography (CT). However, current RMPs are highly simplified and do not exhibit realistic tissue structures or deformation patterns. With the rise of more complex motion compensation technologies such as deep learning-based algorithms, there is a need for more realistic RMPs. This work introduces PixelPrint^4D^, a 3D printing method designed to fabricate lifelike, patient-specific deformable lung phantoms for CT imaging. The phantom demonstrated accurate replication of patient lung structures, textures, and attenuation profiles. Furthermore, it exhibited accurate nonrigid deformations, volume changes, and attenuation changes under compression. PixelPrint^4D^ enables the production of highly realistic RMPs, surpassing existing models to offer more robust testing environments for a diverse array of novel CT technologies.

## 1. Introduction

All forms of in-vivo medical imaging are susceptible to the effects of patient motion, which can degrade diagnostic image quality and impair the localization of structures. Respiratory motion is particularly ubiquitous and has a significant influence on everyday clinical workflows in diagnostic imaging and radiation therapy. Movements that occur during a scan can cause motion artifacts which may obscure critical structures or imitate disease states, leading to misdiagnoses^1^. In radiation therapy, tumor and organ motion is a significant challenge^2,3^, particularly for tumors located in or near the lungs. It can modify dose distribution during therapy, causing inadequate radiation to tumor cells and increased damage to surrounding healthy tissue^4^. This may ultimately result in injury or tumor recurrence.

Many technologies for mitigating the effects of respiratory motion have been and are continuing to be developed^5^. Some well-established radiation therapy motion management strategies include four-dimensional computed tomography (4DCT) for treatment planning, and intensity modulated radiation therapy (IMRT). These allow for the modulation of the radiation beam to better target the expected trajectory of tumor motion. Newer technologies have also been developed, including deformable image registration (DIR), deep learning-based lung tumor prediction models^6,7^, motion artifact reduction algorithms^8,9^, and CT ventilation imaging^10,11^.

These technologies need to be assessed for efficacy both during development and during ongoing use^12^. While clinical trials are necessary for evaluation in real patient scenarios, they have certain limitations such as radiation exposure to patients and a lack of ground truth. Therefore, CT respiratory motion phantoms (RMPs) have been developed to serve as model testing environments. CT imaging phantoms are non-living objects which are imaged instead of patients in the testing, validation, and optimization of various technologies. RMPs are specialized phantoms which can mimic respiratory motion. Thus, RMPs allow researchers to iterate on new technologies and protocols well in advance of patient studies, which facilitates more efficient innovation. They also function as quality assurance tools in the routine maintenance of various equipment.

With the rise of deep learning based respiratory motion management technologies, which are by nature nonlinear and object dependent^13^, there is an increasing need for lifelike “super phantom”^14^ RMPs to serve as accurate and reliable testing environments. Technologies such as CT ventilation imaging rely on precise deformable vector fields (DVFs) and changes in density measured by CT attenuation to assess lung functionality^10,11,15^. Thus, it is crucial that an RMP for assessing CT ventilation include phantom lung structures which can exhibit realistic deformation patterns and attenuation changes. Furthermore, due to significant motion variations between patients^2^, RMPs with patient-specific motion profiles are necessary to evaluate how robust technologies are to these variations.

Many existing RMPs are either rigid phantoms which undergo bulk translation^9,16^ or contain separate rigid portions that slide relative to each other^17–19^. While many of these rigid phantoms have advanced software capable of producing programmable breathing motion paths and patterns, they are not capable of replicating the complex nonrigid deformations which are seen in real patient lungs. Several compressible or inflatable RMPs have been designed to address the nonrigid deformation and attenuation change components of physiological lung motion. Many contain foam structures with high density embedded nodules to mimic tumors^20–28^. A few even include embedded vasculature and airway-mimicking structures^29,30^. Nevertheless, all these RMPs are heavily simplified, with only some major structures included. So far, RMPs made using cadaveric animal lungs contain the most detailed internal structures^31,32^. However, these cadaver phantoms cannot be preserved for long periods due to tissue degradation and thus are not capable of repeatable long-term testing^33^. To our knowledge, no prior RMP exists which includes anatomically accurate and detailed lung structures, can undergo non-rigid deformations, and is also suitable for long term use.

In response to these needs, this study introduces PixelPrint^4D^, a method for 3-dimensional (3D) printing of patient-specific deformable lung phantoms, designed for CT technology development and applicable to 4DCT and respiratory motion management, among other uses. The four dimensions refer to the three spatial dimensions plus one dimension of time. By leveraging components of the previously introduced PixelPrint method, which enables creating rigid lifelike patient-based phantoms^34–41^, PixelPrint^4D^ now fabricates deformable phantoms. PixelPrint converts the attenuation values, measured in Hounsfield Units (HU) from each voxel of patient CT imaging data into fused deposition modeling (FDM) 3D printer instructions called g-code. The target HU value is correlated with printed material density, or infill percentage. The printed material is laid down layer by layer in a line pattern where the line thickness is constantly adjusted to modulate the effective density. If the scale of the line pattern is smaller than the CT detector resolution, the lines will be resolved as regions of varying HU due to the partial volume effect. Past PixelPrint phantoms have demonstrated high levels of structural and HU accuracy, with lung structure measurements of less than a pixel size error and contrast differences of <15 HU compared to the reference patient image^34^. A reader study with five radiologists showed that there was no significant difference in diagnostic and image quality assessments between PixelPrint phantom images and reference patient images^37^. Furthermore, PixelPrint has previously been used in the evaluation of deep learning based CT reconstruction^39^, demonstrating the value of lifelike phantoms for the assessment of complex nonlinear technologies.

In this study we developed a novel method to fabricate patient-specific deformable lung phantoms. The use of a flexible 3D printing material enabled the creation of variable densities and stiffnesses within the phantom. The PixelPrint^4D^ phantom demonstrated lifelike structures and textures as well as realistic DVFs and HU changes when compressed to match patient respiratory phases. By providing a more realistic testing environment, a more robust assessment of CT technologies, especially regarding respiratory motion compensation, can be achieved, ultimately improving the efficiency and quality of their clinical translation.

## 2. Results

### 2.1 Uncompressed lung phantom

The deformable lung phantom was designed based on a respiratory gated chest 4DCT of a radiation therapy patient. The tumor-containing right lung at end inhalation (EI) was selected as the model. Since the phantom was fabricated at EI which is the maximum volume state, all other respiratory states were replicated by compression. A flexible thermoplastic polyurethane (TPU) 3D printing material was selected (Ninjaflex (85A) (Ninjatek, Fenner Precision Polymers, PA, USA)), and a calibration phantom was designed to determine the relationship between 3D printed infill percentage and the attenuation value measured on CT (**Methods – Fig. 6, Methods – Equation 1**). Then the attenuation information within the right lung was converted voxel by voxel into infill percentages according to this calibration equation and subsequently converted to 3D printer instructions using the PixelPrint software^34^. The phantom was 3D printed at full scale using an FDM printer (Lulzbot Taz Sidekick 747 with M175 tool head (Fargo Additive Manufacturing Equipment 3D, LLC, ND, USA)) with TPU (**Fig. 1A**).

**Fig. 1.**
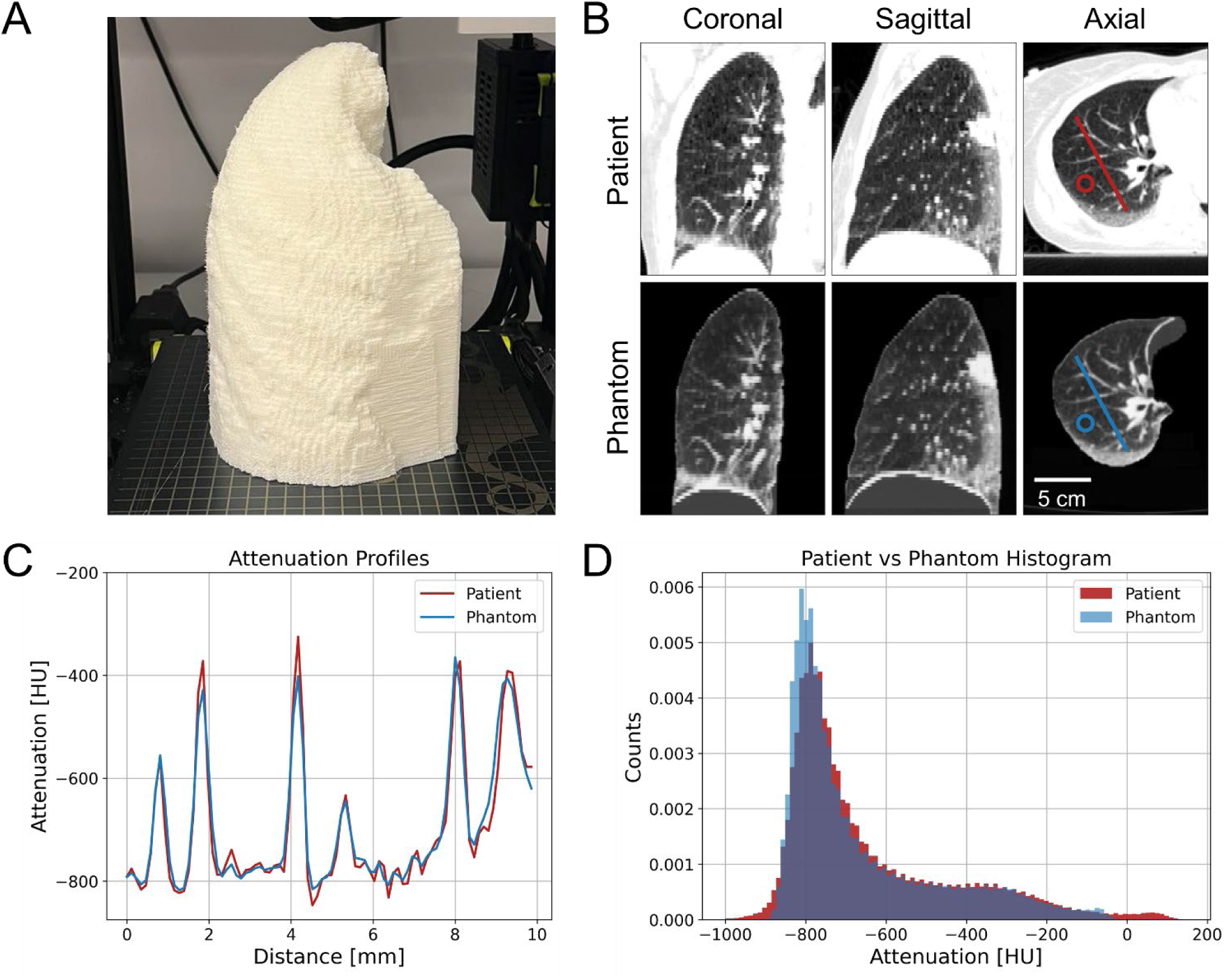
Comparing uncompressed phantom to patient reference image. (A) Completed PixelPrint lung phantom 3D printed out of TPU. (B) CT scan images of the patient and phantom, shown in three orthogonal views. The axial view shows the line where attenuation profiles were measured and circular ROIs where noise was measured. (WL: -500, WW: 1000) (C) Attenuation profiles measured along the lines indicated in (B) on the patient and phantom images (D) Histograms of the patient and phantom images masked to only include regions inside of the lung volume.

A conventional CT scan of the printed phantom clearly replicated structures seen in the patient lung image, including the tumor, bronchi, blood vessels, and even fine details such as lung fissures (**Fig. 1**). Attenuation values were measured along the lines indicated on the axial view of **Fig. 1B** in patient and phantom images, and the resulting attenuation profiles were plotted in **Fig. 1C**. The mean absolute error along the measured lines was 20 ± 19 HU. The mean absolute attenuation error in the background lung parenchyma (where HU < -700) was 28 ± 28 HU and the mean absolute error in the total lung volume was 50 ± 64 HU. Errors in HU were most pronounced in higher density regions such as the tumor and vessels since these regions had higher attenuation values (100 HU) than the maximum attenuation in the phantom (-48 HU). For context, the noise was measured as 45 HU in a relatively homogeneous region of background lung parenchyma in the patient (circular regions of interest (ROIs) in **Fig. 1B**). In addition, the attenuations of the full patient and phantom lungs were plotted as histograms (**Fig. 1D**) and compared using a two sample Kolmogorov-Smirnov (KS) test. Although there was a statistically significant difference when considering the full range of attenuations found in the patient lung (KS-test *p*-value = 0.02 < 0.05), the difference was primarily due to the limited HU range in the phantom. The patient histograms showed an attenuation range that extended 150 HU lower and higher than the attenuation values achieved in the phantom (-840 to -48 HU). Within the attenuation range achieved in the phantom, the histograms showed close overlap, with a KS-test *p*-value of 0.28 > 0.05, indicating no statistically significant difference. Finally, the structural similarity index (SSIM) was measured as 0.93 between the patient and phantom lung, suggesting a high level of structural accuracy in the phantom images.

### 2.3 Compressed lung phantom vs patient 4DCT

Since the phantom was printed in the EI phase, or at maximum lung volume, all other phases from the patient 4DCT were replicated by compressing the phantom. In physiological breathing, the volume of the lung is controlled primarily by the superior/inferior (SI) motion of the diaphragm. In addition, the chest wall also expands and contracts in the anterior/posterior (AP) and, slightly less so, in the medial/lateral (ML) directions to further facilitate lung expansion and contraction. It has been reported that most lung lesions are displaced most significantly in the SI and AP directions^3^. Thus, for this study, a linear compression device capable of compressing the phantom in the SI and AP directions was built (**Fig. 2**). The SI compressor was a 3D-printed clamp-like structure with a moving wall controlled by a lead screw, and a stationary wall (**Fig. 2A**). The lung phantom was positioned between these walls with the diaphragm against the moving wall and the top of the lung against the stationary wall. AP compression was facilitated by a pair of 3D-printed molds placed over the anterior and posterior of the phantom, forming an inverse of the lung shape (**Fig. 2B**). The distance between the two molds was adjusted by tightening screws connecting the two pieces at each corner. The displacements of the diaphragm and chest wall were measured on the patient 4DCT at each phase, and these values then dictated how much the moving wall was displaced for SI compression and how much the AP molds were tightened down for AP compression (**Methods - Table 2**). For this patient, diaphragm displacements ranged from 0 to 15 mm in the SI direction, and the anterior chest wall displacements ranged from -0.6 to 2.2 mm in the AP direction.

**Fig. 2.**
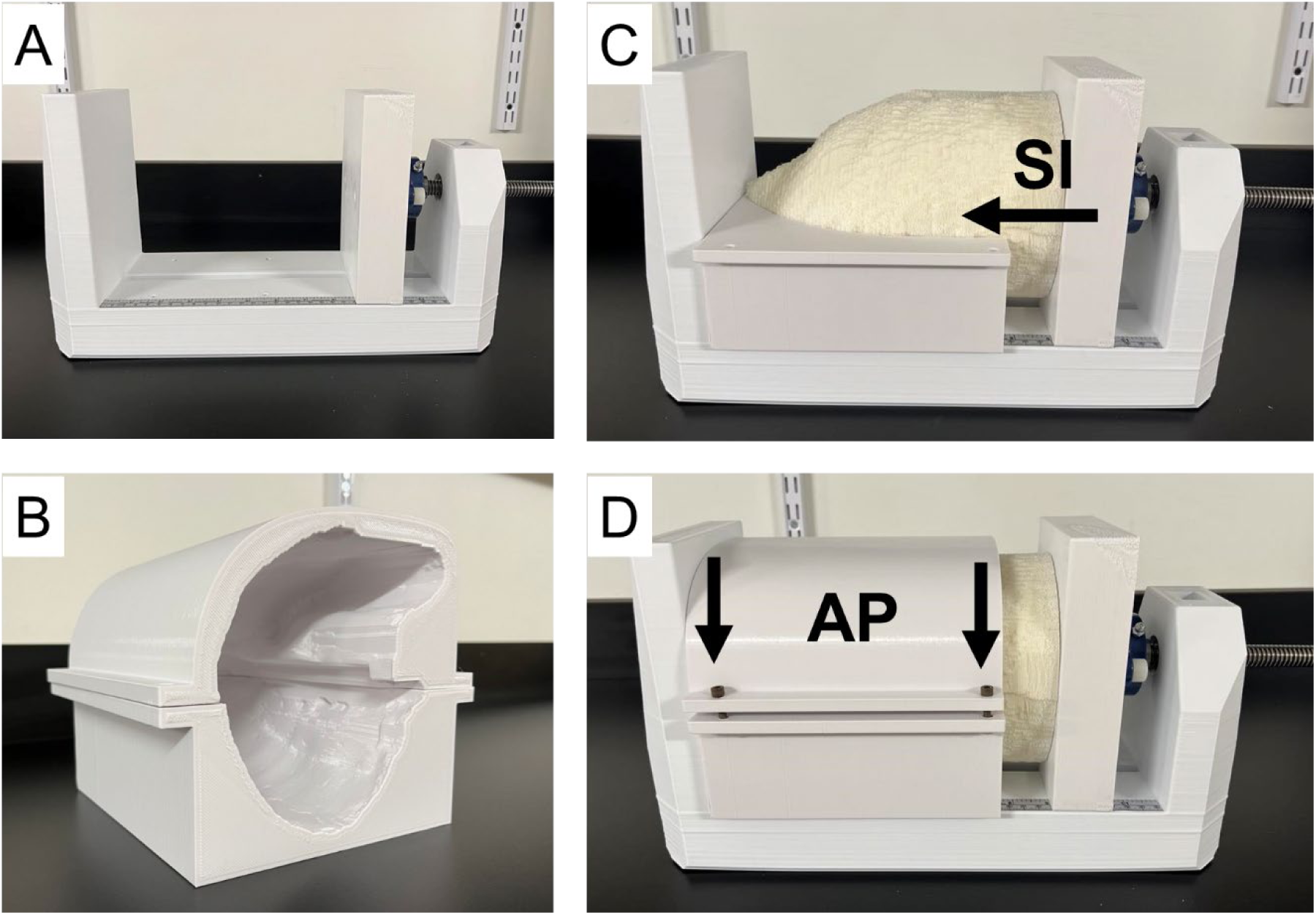
Completed lung phantom with compression device. (A) SI compression device with stationary wall on the left and moving wall on the right. (B) AP compression device with anterior (top) and posterior (bottom) portions (C) Lung phantom inserted into compression device with anterior portion of AP compressor removed to show lung phantom. Arrow indicates direction of movement of moving wall. (D) Full compression device assembled with SI and AP components and lung phantom inserted. Arrow indicates direction of movement of anterior cap.

#### 2.3.1 Pseudo-4DCT

The phantom was imaged with CT at each compression level corresponding to the diaphragm and chest wall displacements measured from the patient 4DCT (**Fig 4A**). The resulting series of phantom images were combined to form a pseudo-4DCT. The phantom pseudo-4DCT and patient 4DCT were then stitched together into an image sequence and looped to form a video showing the motion of the phantom and patient lung side by side during the respiratory cycle. This video can be made available upon request.

#### 2.3.2 Displacements

To assess the deformation characteristics of the phantom, DIR^42^ was performed on the phantom compression images and patient 4DCT. This aligns all structures from image to image and allows for direct comparisons of a given location in the lung between phases. The DIR algorithm also yielded a set of deformation vector fields (DVFs) which represent the local displacements of each location throughout the lung volume relative to EI (respiratory phase 0%). The phantom results were then compared to the patient (**Fig 3A**).

**Fig. 3.**
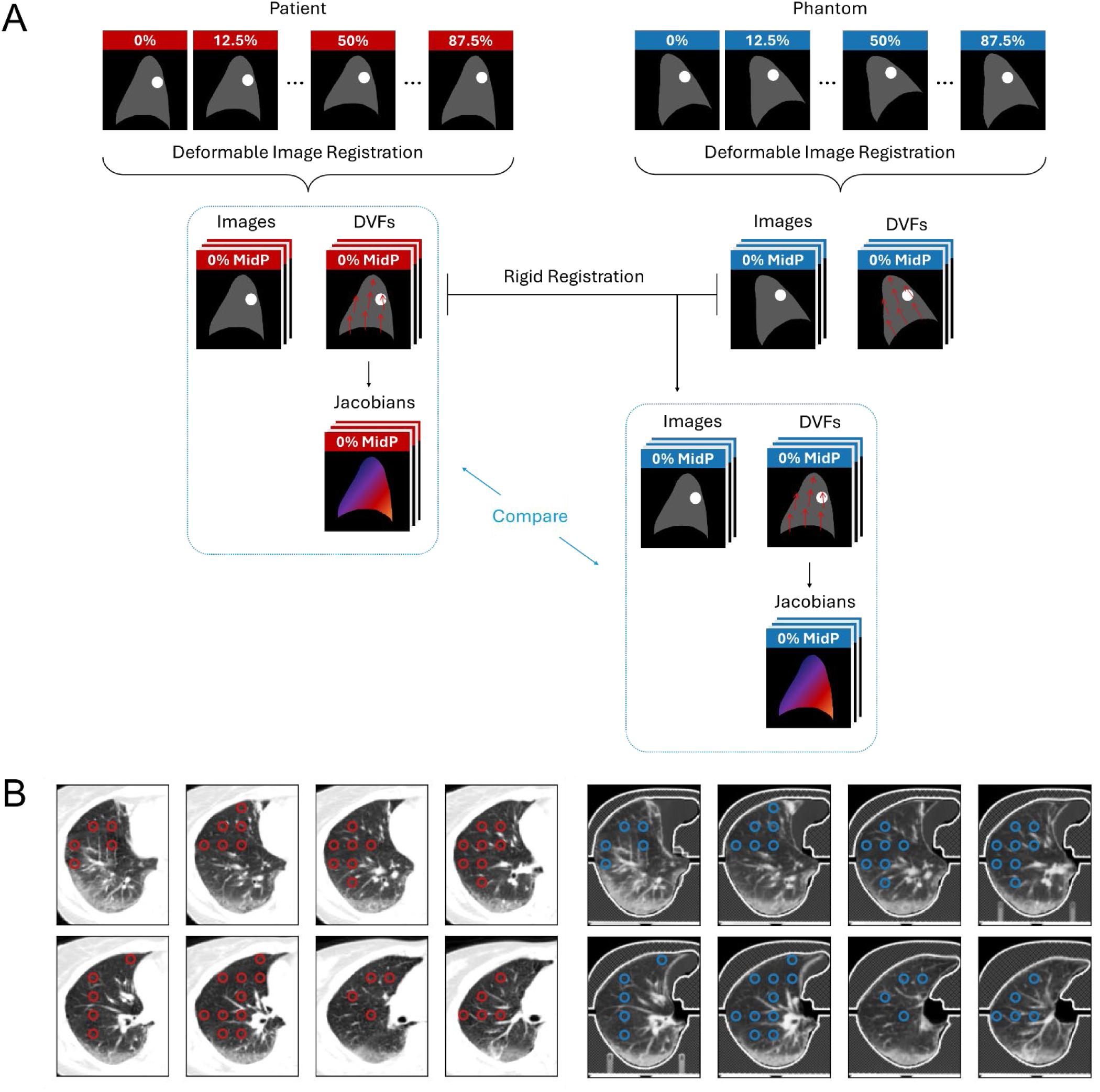
Image analysis pipeline for assessing phantom deformation characteristics. (A) Image processing steps starting from patient 4DCT (left), and images of the phantom (right) compressed to match each phase of the patient 4DCT. DIR and rigid registration were performed to obtain aligned CT image volumes, DVFs, and Jacobian maps which were compared to assess the accuracy of the phantom deformations. (B) Fifty ROIs selected in the background lung parenchyma where mean attenuation and Jacobian values were measured over the course of the respiratory cycle.

To assess the accuracy of the phantom tumor motion, displacements of the tumor were measured in both the patient and phantom over the course of the respiratory cycle (**Fig. 4C**). The values of the DVF were measured at the center of the tumor in each image. The mean tumor motion errors defined as the phantom displacements minus patient displacements were 0.7 ± 0.6, -0.4 ± 0.5, and 0.1 ± 0.4 mm in the SI, AP, and ML directions respectively. Positive values indicate that the phantom displacements are greater in the superior, anterior, and medial directions compared to the patient, and vice versa. Notably, these mean errors are each smaller than the voxel size of the patient image in the corresponding axes (3.00 x 1.17 x 1.17 mm).

**Fig. 4.**
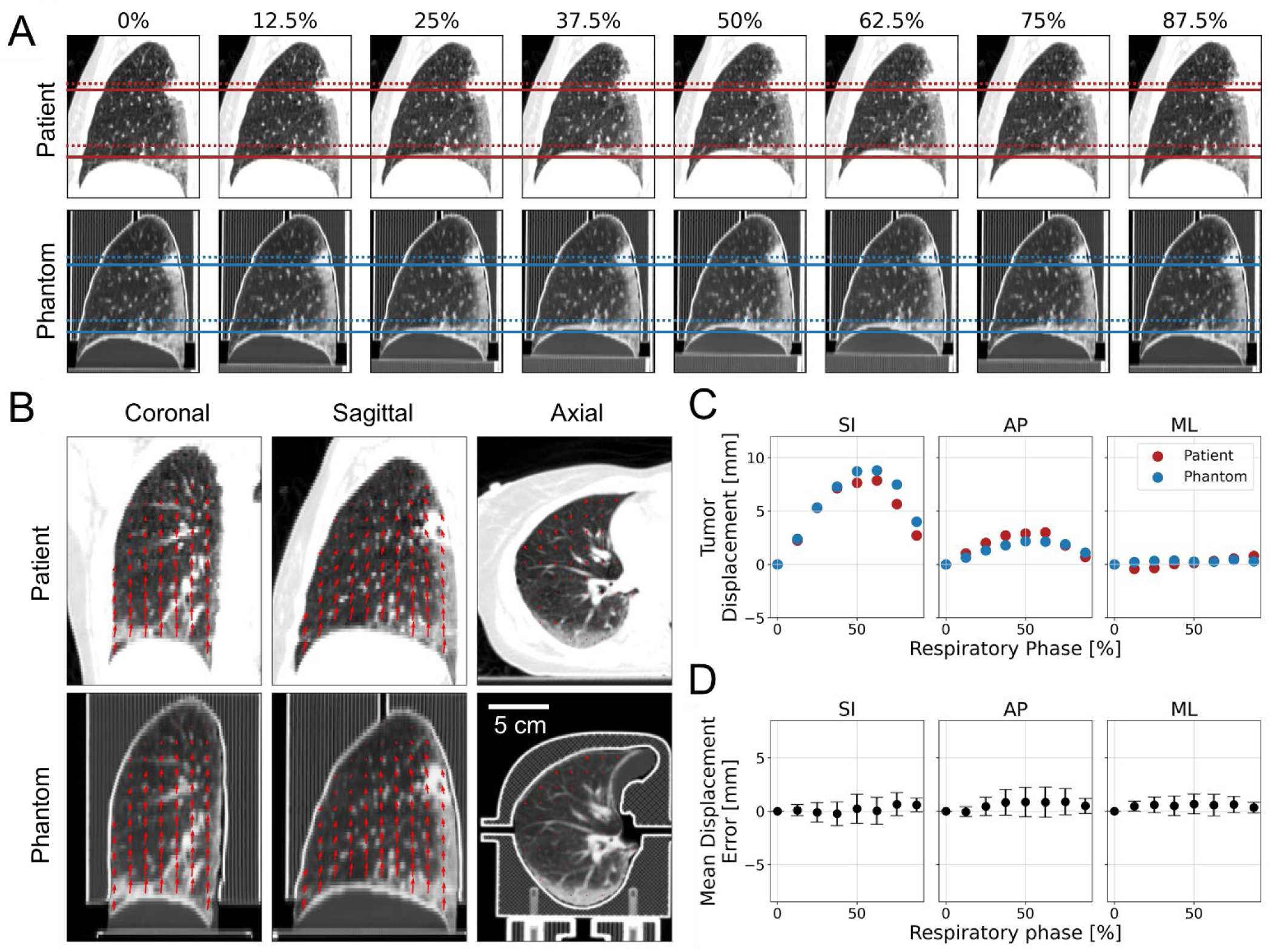
Displacements of the phantom and patient lungs. (A) Sagittal views of each phase from the patient 4DCT and matched phantom compression scans showing motion of the patient and phantom lung during the respiratory cycle. The SI position of the tumor center and top of the diaphragm are shown with dotted and solid lines at EE (62.5%) and EI (0%) respectively (B) DVFs at the 62.5% respiratory phase displayed as a field of arrows overlaid on the patient and phantom CT images, shown in three orthogonal views. The arrow directions correspond to the vector directions and the lengths are proportional to the magnitude of displacement (WL: -500, WW: 1000). (C) Displacements of the patient and phantom tumor over the course of the respiratory cycle, measured in each orthogonal direction. Positive values correspond to displacements in the superior, anterior, and medial directions. (D) Mean displacement error of the phantom DVFs compared to the patient throughout the total lung volume.

While tumor displacements are the most important motion to consider for radiation therapy treatment planning, it is also valuable to quantify the motion of other structures in the lung to help minimize damage to healthy surrounding structures. Therefore, we also assess the phantom’s ability to replicate non-rigid deformations of non-tumor lung structures by comparing the DVFs throughout the whole patient and phantom lung volumes. The DVFs at the end exhale (EE) phase (respiratory phase 62.5%) are shown in **Fig. 4B**, demonstrating variable deformations throughout the lung volume. The phantom DVFs throughout the lung at each phase are shown in **Fig. 5D**. At EE the mean errors were 0.0 ± 1.3, 0.8 ± 1.4, and 0.6 ± 1.0 mm in the SI, AP, and ML directions respectively (**Fig 5D**). Positive values indicate that the phantom displacements are greater in the superior, anterior, and medial directions compared to the patient, and vice versa. Again, these mean errors are smaller than the voxel size of the patient image in the corresponding axes.

**Fig. 5.**
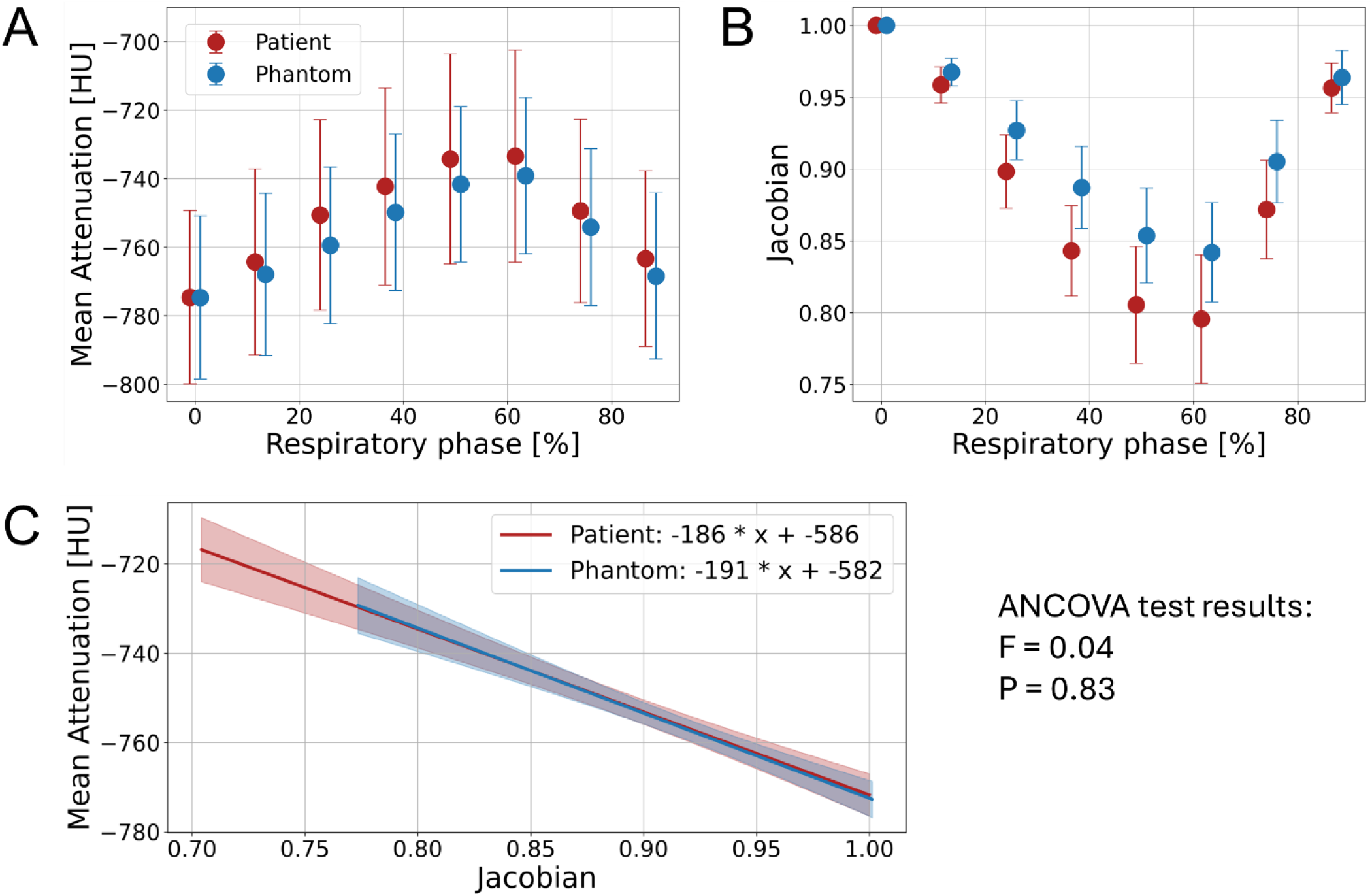
Attenuation changes and volume changes. (A) Mean attenuation measured in the background lung parenchyma across different phases in the patient and matched phantom compression scans. (B) Mean Jacobian values representing volumetric changes measured in the background lung parenchyma across different phases in the patient and matched phantom compression scans. (C) The linear regression relating attenuation and Jacobian in the patient and phantom, along with the 95% confidence interval indicated with the shaded regions.

#### 2.3.3 Attenuation and volume changes

Next the compression in the background lung parenchyma (< -700 HU) was assessed by measuring attenuation changes and local volume changes throughout the respiratory phases (**Fig 5**). These quantities are used in CT ventilation imaging to assess lung functionality, where greater changes in attenuation and volume during respiration are assumed to indicate more effective lung function. In both the patient and phantom, the mean attenuation was found to increase during expiration/compression and decrease again during inspiration/decompression (**Fig 5A**). The maximum attenuation increases relative to the EI state were 41 ± 22 HU in the patient and 36 ± 12 HU in the phantom, each measured at EE. This was a mean attenuation change error of 5 HU, or 12%.

Local volume changes can be estimated by using Jacobians, which were calculated from the DVFs (Equation (3))^43^. Jacobian = 1 corresponds to no volume change, while values < 1 indicate volumetric contraction and values > 1 indicate volumetric expansion. In this study, all Jacobians were calculated relative to the EI state, so all values are ≤ 1. An increase in Jacobian indicates relative volume expansion between phases and vice versa. The mean Jacobians were measured in the same locations where the attenuations were measured in the background lung parenchyma (**Fig 5B**). In both the patient and phantom image series, the lung volumes decreased during expiration/compression. The greatest compression was measured at EE, with Jacobian = 0.80 ± 0.04 and 0.84 ± 0.03 in the patient and phantom respectively. This corresponds to volumetric contractions of 20% and 16% in the patient and phantom, or an error of 20%.

Finally, the relationship between attenuation and volume changes was assessed by performing a linear regression between the mean attenuations and Jacobian measures (**Fig 5C**). The regressions for the patient and phantom had a strong correlation, with a 2.7% difference in slope and 0.7% difference in intercept. Analysis of covariance yielded *p* = 0.83 and *f* = 0.04, which indicated that there is no significant difference between the two regressions. This suggests that the phantom behaves realistically in terms of its relationship between volume change and attenuation change.

## 3. Discussion

We have developed PixelPrint^4D^, a method of fabricating patient-based deformable lung phantoms for CT imaging with realistic lung structures that exhibit lifelike deformation patterns. PixelPrint^4D^ phantoms have the potential to serve as low-cost lifelike testing environments for various CT respiratory motion compensation technologies including tumor tracking and CT ventilation imaging. The lung phantom in this study demonstrated an SSIM of 0.93 compared to the reference patient lung image, showing high structural accuracy. When compressed in the SI and AP directions to match the diaphragm and chest wall displacements from the patient 4DCT, the phantom showed similar deformation patterns to the patient. Phantom tumor displacements and DVFs throughout the lung matched the patient tumor displacements in all directions with mean errors within the size of one voxel (3.00 x 1.17 x 1.17 mm) of the patient CT slices. The phantom also demonstrated attenuation changes within 5 HU of the patient, and percent volume changes within 4% of the patient.

To our knowledge there are no other deformable lung phantoms which display the level of accurate internal lung details and deformation patterns found in the PixelPrint^4D^ phantom. The most common RMPs are made of rigid components, which cannot replicate the non-uniform deformations found in patient lungs. While some previous RMPs have included some vascular or airway structures^29,30^, they do not exhibit as much detail nor include realistic textures in the background lung parenchyma as the PixelPrint^4D^ lung phantom does. In both structure and function, PixelPrint^4D^ surpasses the current state of the art of RMPs.

A recent publication has highlighted the importance of closing the gap between the current highly simplified phantoms and the much more complex human tissues, especially given the rapid advancement in medical imaging technologies^14^. We believe PixelPrint^4D^ represents a major step in this direction. The high level of realistic detail in PixelPrint^4D^ will be especially advantageous in the assessment of more complex applications. For example, deep learning-based algorithms have nonlinear and object dependent performance^13^, which can produce different results between testing with a simplified non-realistic phantom and in real patients. Another example is CT ventilation imaging which utilizes nonrigid deformations and attenuation changes. Previous RMPs would not be suitable for this application, but this study demonstrated that PixelPrint^4D^ is, since it closely imitates the nonrigid deformations and attenuation changes seen on patient imaging. Additionally, this technology facilitates the printing of models representing various patients with different diseases or specific technical targets, helping to build a library of phantoms for various applications. Furthermore, since the 3D-printing fabrication process described in this study is highly customizable, PixelPrint^4D^ has the potential to be adapted for additional use cases, such as by adding dosimetry devices to measure dose distributions.

There are several limitations of the current study. First, the compression system used for testing the lung phantom only enabled step-by-step compression rather than continuous dynamic motion. A dynamic compression system capable of continuous real-time motion will be essential for testing technologies such as 4DCT. This will furthermore enable experiments assessing time-dependent compression characteristics of the phantom such as hysteresis. Another limitation is that the maximum HU value attainable in the current design was -48 HU at 0% compression, which is about 150 HU lower than maximum HU in the patient lung. Future investigations into other printing materials or printing parameters may improve the phantom attenuation range to better match clinical attenuation values. In addition, the current motion system only enables SI and AP compression and not ML compression, which is consistent with the current standards in the field. However, ML motion can have a significant contribution in certain cases^3^ and thus would be valuable to incorporate in future studies. Finally, this study focused on the design and development of a single lung phantom and did not yet include other surrounding structures such as the torso, ribs, heart, and the other lung. Including these other structures would provide more realistic attenuation profiles surrounding the lungs, which is important, for example, when studying dose distribution effects. In the future, we plan to design a comprehensive motion system which includes these other structures and is capable of dynamic programmable motion.

In conclusion, the PixelPrint^4D^ deformable lung phantom developed in this study exhibits realistic lung structures and deformation characteristics on CT imaging, making this phantom a valuable tool for assessing motion effects on CT lung imaging. PixelPrint^4D^ offers a distinct environment that can expedite and solidify advancements in CT, especially in respiratory motion compensation technologies, compared to traditional CT RMPs, leading to better patient care and treatment.

## 4. Methods

### 4.1 PixelPrint process

PixelPrint utilizes 3D-printers and lays down material at varying speeds to modulate the density and thus the CT attenuation of the phantom on a voxel-by-voxel basis. In this work, all phantoms were 3D printed using an FDM printer (Lulzbot Taz Sidekick 747 M175 tool head, Fargo Additive Manufacturing Equipment 3D, LLC, ND, USA) which feeds heated material in the form of filaments through a nozzle (E3D V6 0.25 mm brass) to lay down material in multiple 2D layers. These 2D layers stacked together form a 3D object. Each 2D layer in a PixelPrint phantom consists of parallel lines with fixed spacing but variable line widths. The width of the line is modulated continuously by changing the speed at which the printing nozzle moves while keeping the extrusion rate of the filament fixed. This results in a partial volume effect where different ratios of filament and air within a single imaging voxel appear as different attenuation levels on CT imaging. The percentage of volume filled by the 3D printing material is called the infill percentage. PixelPrint utilizes this effect to map attenuation values in each voxel of a patient CT image to the target 3D-printed infill percentage, thus reproducing highly detailed features from the reference patient image. More information on the PixelPrint process can be found in the original publications^34,38^.

### 4.2 Flexible 3D-printing materials selection

PixelPrint originally used polylactic acid (PLA), a rigid plastic 3D printing material. To fabricate a deformable phantom, several flexible 3D printing filaments were assessed for compatibility with PixelPrint technology. To emulate human lung tissue with attenuations of about -800 HU, the material must be printed with low infill percentages of 20% or less. We targeted a line spacing 1 mm, or roughly the pixel size of standard 4DCT scans, and a layer height of 0.2 mm which is smaller than the slice thickness of most standard CT protocols. Thus, for an infill of 20% or less, the minimum line thickness needed to be less than 0.2 mm for materials with similar density to PLA^34^. Several materials were assessed including Ninjaflex (85A) (Ninjatek, Fenner Precision Polymers, PA, USA), Chinchilla (Ninjatek, Fenner Precision Polymers, PA, USA), Caverna ST (Infinite Material Solutions, LLC, WI, USA), Polyflex TPU90 (Polymaker, LLC, TX, USA), and Filaflex 85A (Recreus, Alicante, Spain). We use each of these materials to print test phantoms with infills between 15% and 100% using the PixelPrint method. Many of these materials were prone to have breaks in the printed lines and/or form clumps, especially in the lowest infill regions of 15%. This resulted in uneven attenuations on CT images. Caverna ST, although it did not have issues with print quality, required a washing step to dissolve part of the material, which led to uneven washout throughout the phantom and uneven attenuation values on CT. Of the materials tested, Ninjaflex, a type of thermoplastic polyurethane (TPU), at a filament diameter of 1.75 mm produced the most reliable and consistent print quality at infills from 15% to 100% and was selected for this phantom. Ninjaflex TPU filament had a shore hardness of 85A, a specific gravity of 1.19 g/cc, a tensile yield strength of 4MPa, and 65% elongation at yield (https://ninjatek.com/shop/ninjaflex/).

### 4.3 HU calibration

Next a calibration phantom was printed to determine the relationship between printed infill percentage and CT attenuation for TPU. The calibration phantom was designed as a 10 cm diameter by 1 cm high cylinder and consisted of 10 homogeneous wedges with infills of 15%, and 20-100% at 10% increments (**Fig. 6**). It was printed on an FDM printer (Lulzbot Taz Sidekick 747 with M175 tool head) with a 0.25 mm brass nozzle (E3D V6). The nozzle temperature was set to 220°C and the bed temperature was set to 40 °C. With the extrusion rate set to 0.6 mm^3^/s, a layer height of 0.2 mm, and line spacing of 1 mm, the nozzle speeds needed to obtain 15-100% infills were between 20 and 3 mm/s. Total print time was 1 day. The calibration phantom was imaged with a CT scanner (Spectral CT 7500, Philips Healthcare, MA, USA) with scan and reconstruction parameters listed in Table 1. Mean attenuation was measured in 13 mm diameter circular ROIs placed in the center of each wedge. A linear regression was performed between the mean attenuation and the infill percentage of each wedge to determine the TPU calibration function (**Fig. 6C**).

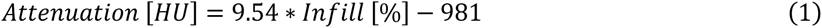

**Fig. 6.**
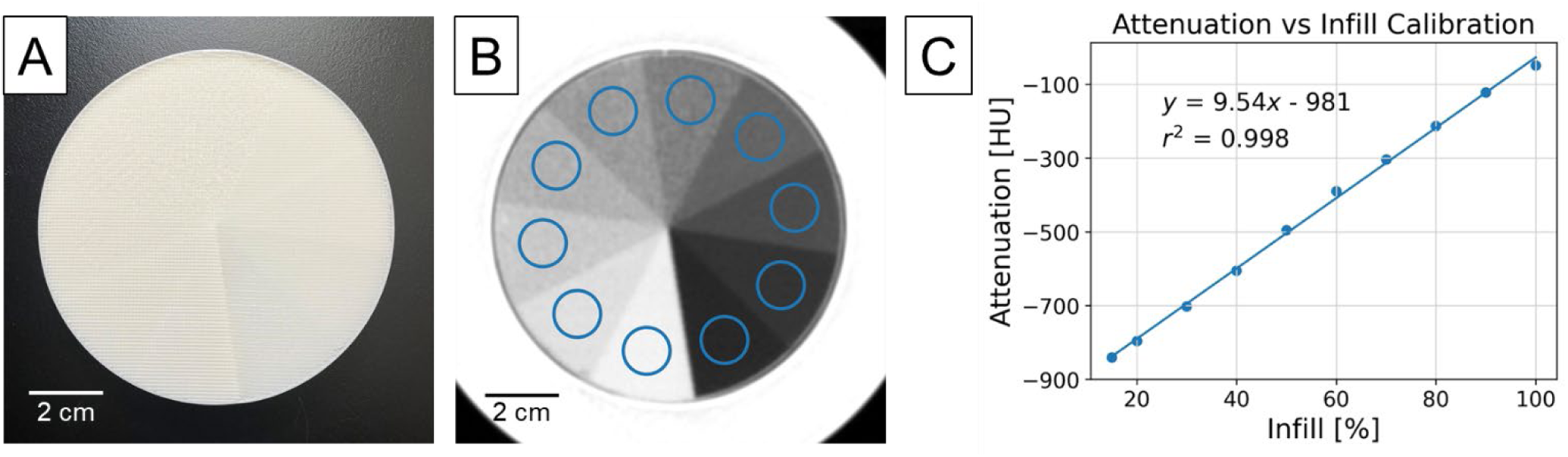
Calibration phantom. (A) Calibration phantom 3D printed out of TPU using the PixelPrint method. (B) CT scan image of the calibration phantom with regions of interest (ROIs) showing where attenuation was measured for each wedge (WL: -500, WW: 1000) (C) Measured attenuation values plotted against the 3D printed infill percentage, along with the linear regression which served as the calibration function.

**Table 1.** CT scan parameters.

|  | Calibration phantom scans | Patient 4DCT scan | Lung phantom scans |
| --- | --- | --- | --- |
| **CT Scanner** | Spectral CT 7500<br>(Philips Healthcare) | Gemini TF Big Bore<br>(Philips Healthcare) | Spectral CT 7500<br>(Philips Healthcare) |
| **Tube voltage [kVp]** | 120 | 120 | 120 |
| **Exposure [mAs]** | 150 | 602 | 600 |
| **Scan mode** | Helical | Helical | Helical |
| **Slice thickness [mm]** | 1 | 3 | 1 (no compression)<br>3 (compression) |
| **Field of View [mm]** | 128 | 600 | 300 |
| **Matrix size** | 512 x 512 | 512 x 512 | 512 x 512 |
| **Pixel spacing [mm]** | 0.25 | 1.1719 | 0.5859 |
| **Reconstruction Filter** | B | B | B |

### 4.4 Deformable lung phantom fabrication

This retrospective study was approved by the University of Pennsylvania Institutional Review Board (Protocol # 853697). A respiratory gated chest 4DCT of a radiation oncology patient with a 2.5 cm tumor in the upper right lung lobe was selected as a reference for the fabrication of the deformable lung phantom. The 4DCT included eight respiratory gated phases and the scan parameters are listed in Table 1. The phantom was fabricated based on the image from the end inhale (EI) state, or 0% phase, which represents the maximum lung volume from the 4DCT. The right lung was segmented from this image using an automated deep learning algorithm^44^ and manually adjusted in ITK-SNAP (www.itksnap.org)^45^ to include more of the right main and secondary bronchi. The full segmented right lung, along with about 4 mm of diaphragm directly below the lung, were converted to 3D printer instructions, or g-code, using the PixelPrint software^34^ and TPU calibration function (Equation (1)). The phantom was printed using the same parameters as the calibration phantom with the z-axis of the printer corresponding to the superior/inferior axis of the lung. The region below the lung was filled with a uniform infill of 25% to physically support the phantom while printing, due to the dome shape of the diaphragm. The finished phantom was 23 cm in height, and total printing time of the phantom was two weeks.

### 4.5 Compression device

Since the phantom was initially fabricated in the EI or maximum volume state, the phantom needed to be compressed to achieve the other states in the respiratory cycle. During physiological respiration, muscles around the chest including the diaphragm and intercostal muscles contribute to the expansion and contraction of the lungs. The primary direction of motion is in the superior/inferior (SI) direction^2^ due to the upward and downward motion of the diaphragm. To emulate the motion of the diaphragm, a linear compression device was designed, and consisted of a holder with a vertical stationary wall, and a moving wall controlled by a lead screw assembly. The compression device was designed in a computer aided design (CAD) software (Inventor Professional, Autodesk, CA, USA). The holder and its vertical wall were 3D printed out of polyethylene terephthalate glycol (PETG) and the moving wall was 3D printed out of PLA using FDM printers (Prusa XL for the holder and Lulzbot Taz Sidekick 747 with M175 tool head for the moving wall).

Additionally, an anterior/posterior (AP) compression apparatus was designed to induce compression along the AP axis to mimic the motion of the chest wall. The AP compression apparatus was designed as a pair of molds which fit the shape of the anterior half and posterior half of the lung phantom. The segmentation of the patient lung was dilated by 2 mm to allow for tolerance, and cutouts were added to ensure that the lung phantom would be able to slide into the molds. This adjusted segmentation was then inverted and converted into a standard triangle language (STL) file using the marching cubes algorithm (scikit-image.measure toolkit). In a CAD software (Fusion 360, Autodesk, CA, USA), the inverted mold was separated into an anterior portion and a posterior portion with a 6 mm gap in between to allow for compression. Mesh smoothing was applied to ensure a smooth internal surface for reducing friction between the AP compression device and the phantom Extension wings were added to the medial and lateral sides of the parts to allow the parts to be attached by screws at the four corners. Both portions were 3D-printed on FDM printers (Lulzbot Taz Sidekick 747 with M175 tool head and Lulzbot Taz6) using PLA. The level of compression was adjusted by tightening and loosening the attachment screws to change the spacing between the parts.

### 4.6 Lung phantom image acquisition

The phantom was first scanned in its uncompressed state to assess the quality of the phantom print before applying any deformation. The lung phantom scan protocols were designed to match the patient scan protocols with a few exceptions. This scan was performed with a slice thickness of 1 mm, which is a higher resolution than the input patient image (Table 1). A smaller slice thickness was selected since a scan with thicker slices might not line up with the same slices in the patient image. Information from adjacent patient image slices would then be averaged into one slice in the phantom, thus preventing direct slice to slice comparisons between the patient and phantom. A smaller slice thickness mitigates this issue. In addition, the phantom was scanned at a field of view (FOV) of 300 mm, or half that of the patient (600 mm), since the patient FOV was larger than the maximum FOV setting on the CT scanner used.

Then the phantom was scanned inside the compression device at varying states of compression. The phantom was inserted into the AP compression apparatus which was then inserted into the larger SI compression device, thus enabling both SI and AP compression. The posterior of the phantom was positioned toward the CT bed while the anterior portion faced upward. The superior portion of the lung phantom was oriented toward the stationary wall and the inferior portion was oriented toward the moving wall. The moving wall was used to compress the phantom in the superior/inferior axis similar to the diaphragm.

The SI displacements of the diaphragm and AP displacements of the anterior chest wall were measured on the patient 4DCT and used to determine the amount of compression applied to the phantom. Since the portion below the phantom diaphragm was printed with a low infill, this region was also compressible. This resulted in the diaphragm moving less than the input motion from the wall. To compensate for this effect such that the phantom diaphragm displacements matched the patient diaphragm displacements, the SI wall was displaced by 1.25x the patient diaphragm displacements. The phantom was simultaneously compressed in the AP direction by tightening the AP compression screws to match the anterior chest wall displacements measured from the patient 4DCT. The phantom was then imaged with CT (Table 1) at each compression level (Table 2). For these compression scans, we matched the slice thickness of the patient images (3 mm).

**Table 2.**
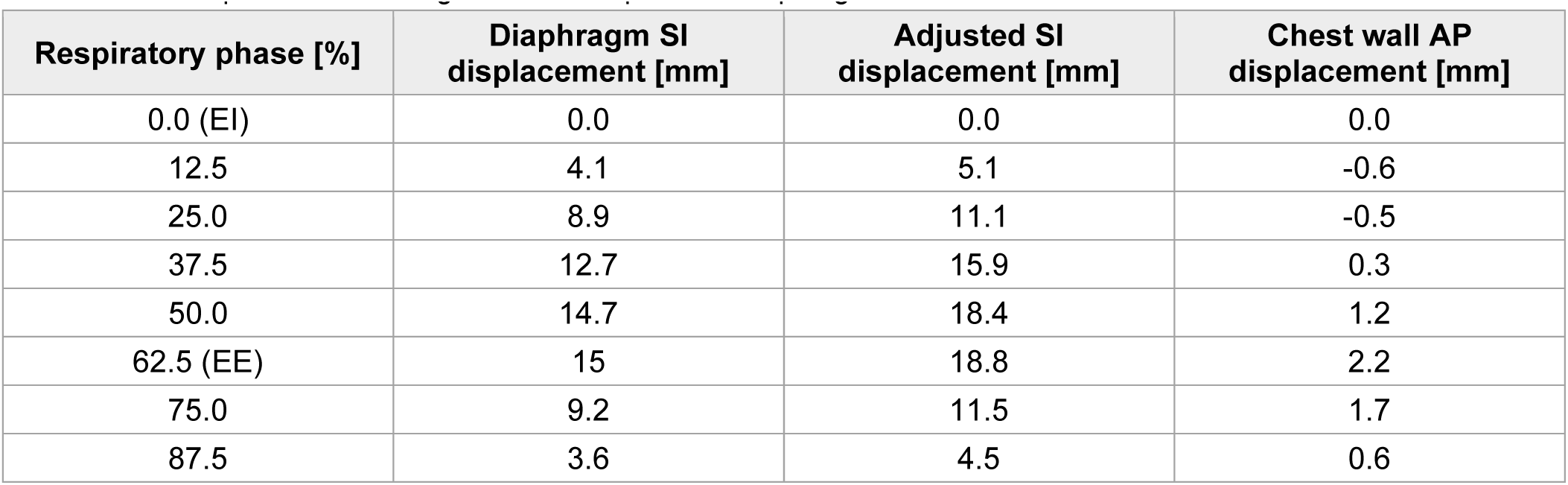
Displacements of the patient diaphragm in the superior direction and of the patient anterior chest wall in the posterior direction. These displacements were used for compressing the lung phantom, where SI displacements were adjusted to account for additional compression in the region below the phantom diaphragm.

| Respiratory phase [%] | Diaphragm SI displacement [mm] | Adjusted SI displacement [mm] | Chest wall AP displacement [mm] |
| --- | --- | --- | --- |
| 0.0 (EI) | 0.0 | 0.0 | 0.0 |
| 12.5 | 4.1 | 5.1 | -0.6 |
| 25.0 | 8.9 | 11.1 | -0.5 |
| 37.5 | 12.7 | 15.9 | 0.3 |
| 50.0 | 14.7 | 18.4 | 1.2 |
| 62.5 (EE) | 15 | 18.8 | 2.2 |
| 75.0 | 9.2 | 11.5 | 1.7 |
| 87.5 | 3.6 | 4.5 | 0.6 |

### 4.7 Comparing patient and uncompressed phantom

First the phantom in uncompressed state was compared to the patient image at EI. The phantom image was rigidly registered to the patient image using ITK-SNAP (www.itksnap.org)^45^ to ensure alignment of images for comparison. The attenuation values were measured along a line passing through regions of background lung parenchyma and vasculature in an axial slice in both the patient and phantom. The mean absolute attenuation error along the attenuation profiles was calculated. The attenuations in the full lung were also assessed by measuring the mean absolute error. This was calculated as the mean of the absolute difference image between the phantom and patient images masked to include only those regions within the right lung segmentation. To assess the errors only in the background lung parenchyma, we defined the background lung as regions with attenuation value < -700 HU. The background mean absolute attenuation error was then calculated as the mean of the absolute difference image masked to only the background. Histograms of the attenuations within the lung segmentation were plotted for the patient and phantom images to compare attenuation distributions. The histograms were compared with a two-sample two-sided Kolmogorov-Smirnov (KS) test (scipy.stats.ks_2samp), both over the full range of attenuations found in the patient lung (-1000 to 370 HU), and within the attenuation range attainable with the current TPU material and printing parameters (-840 to -48 HU). Image noise was measured as the standard deviation of attenuation values in a 1.5 cm diameter circular ROI placed in a relatively homogeneous region of background lung parenchyma. Finally, the SSIM (skimage.metrics.structural_similarity) was computed between the patient and phantom images. To compare only the regions within the lung volume, the images were masked with the right lung segmentation and cropped to the edges of the lung. A normalized gaussian kernel of width sigma=1.5 was used and the data range was set to the attenuation range of the patient lung (-1000 to 370 HU).

### 4.8 Comparing patient 4DCT and phantom pseudo-4DCT

Next the image sequence of the phantom in various compression states were compared to the original patient 4DCT images. The series of phantom images were combined and played in sequence to form a pseudo-4DCT. This, along with the gated images from the patient 4DCT were animated side by side in Slicer3D, an open-source software for visualizing medical data (https://www.slicer.org/), as a video to show how the tumor, vessels, and other lung structures move in space relative to each other over the course of the respiratory cycle (Supplementary Video 1).

### 4.9 Quantitative comparison of patient and phantom deformations

Finally, the deformation characteristics from the images of the compressed phantom were compared quantitatively to the patient 4DCT. To do this, DIR was performed separately on the series of phantom compression scans and on the patient 4DCT using a fast elastic image registration (FEIR) protocol^42^. The first output of DIR was the middle position (MidP) states of the patient and phantom lungs, calculated as the geometric middle of the input images. The second output was the series of patient and phantom images each registered to the MidP, such that all structures are aligned from image to image. This allows for direct comparison of attenuation values from the same region of lung tissue from phase to phase. The third output of the DIR algorithm was a set of DVFs indicating displacements throughout the lungs in each orthogonal direction for each phase with respect to the MidP. The DVFs with respect to the EI state were then calculated as:

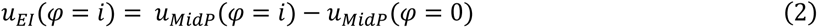

Where *u_EI_* (*φ* = *i*) is the DVF at phase *i* with respect to EI, *u_MidP_* (*φ* = *i*) is the DVF at phase *i* with respect to MidP, and *u_MidP_* (*φ* = 0) is the DVF at phase 0 with respect to MidP. The DVFs with respect to EI were then used to calculate the Jacobian *J*, which quantifies the local volumetric change in each image with respect to the EI state^43^:

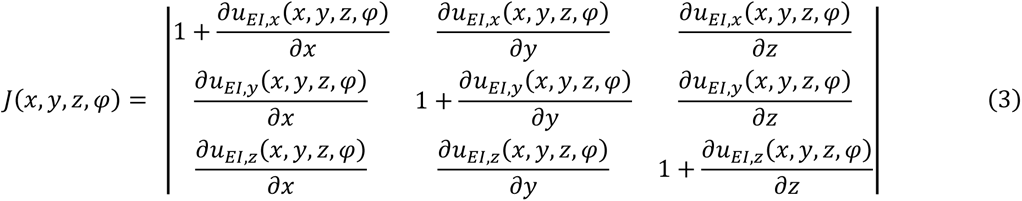

Where *u_EI,x_*, *u_EI,y_*, and *u_EI,z_* are the *x*, *y*, and *z* spatial components of the vector field ***u*,** and (*x,y,z,φ*) refer to the *x*, *y*, and *z* location in the image at phase *φ*. A Jacobian of 1 indicates no change, > 1 indicates local expansion, and < 1 indicates local contraction. After this, the DIR image set, DVFs, and Jacobian maps of the phantom were rigidly registered to the patient series in ITK-SNAP (www.itksnap.org)^45^ to allow direct comparisons between each location in the patient and phantom. Finally the right lung volume was segmented on the DIR patient image using an automated deep learning lung segmentation tool^44^.

We then assessed the accuracy of the phantom tumor motion. The displacements at the center of the phantom and patient tumor were measured from the DVFs and compared, and the mean displacement error of the tumor over the course of the respiratory cycle was calculated for each orthogonal direction (SI, AP, and ML). Since all images have already been registered such that the lung structures are aligned from phase to phase, the center of the tumor only needed to be manually selected once for all images. Additionally, the mean absolute errors in DVFs within the full lung volume at each phase were also computed using the lung segmentation as a mask.

Next, the attenuation changes in the background lung parenchyma were assessed. Fifty 1 cm diameter circular regions of interest (ROIs) (**Fig. 3B**) were selected in the parenchyma of the registered images, defined as regions with mean attenuation < -700 HU. Additionally, regions containing noticeable vasculature or pathology were excluded. Since the images had already been registered, the ROI locations are the same between phases. For each ROI, the mean attenuation and Jacobian values were measured across all phases in the phantom and patient images. Finally, a linear regression assessing the relationship between attenuation changes and Jacobian values was performed for the patient and phantom individually (statsmodels.api)^46^ to obtain linear fits and the 95% confidence interval. These regressions were compared using analysis of covariance (pingouin.ancova)^47^.

## Data Availability

All data produced in the present study are available upon reasonable request to the authors.

## 5. Acknowledgements

The authors would like to thank Dr. Boon-Keng Kevin Teo for selecting the patient 4DCT used as a reference for the phantom in this study and Magdalena Bazalova-Carter for inspirational discussions. We acknowledge support through the National Institutes of Health (R01EB035092, R01EB030494, & R01EB031592) and Philips Healthcare.

## 6. Author contributions

Conception: JI, PN,

Design: JI, NM, KM, MG, PN

Data acquisition: JI

Analysis: JI, AP

Drafting: JI

Revising: JI, KM, PN

